# On the variability of EuroMOMO reporting of all-cause all-ages mortality z-scores

**DOI:** 10.1101/2025.05.08.25327238

**Authors:** Goizalde Badiola-Zabala, Manuel Graña

## Abstract

**Objectives:** Assess the persistence of data reporting in EuroMOMO web site. Detect weeks with high variability due to amendments carried out after the consolidated publication.

**Study design:** Retrospective web examination looking for amendments of weekly all-cause all-ages mortality z-scores published in EuroMOMO site since the year 2018 until october 2023.

**Methods:** Past published all-age all-cause mortality z-scores for all collaborating countries have been recovered monthly from the Internet Archive. The variance of each week reported z-scores is computed accross the recoverd Internet Archive samples. A Poisson distribution fitted to the variance time series allows to detect weeks with significant variance (*p <* 0.01) and extremely significant variance (*p <* 0.001) .

**Results:** Over the entire pool of countries, the study finds 179 weeks with significan variance in the reports, among them 61 weeks of extremely significant variance. France, UK, and Germany have the largest number of these weeks.

**Conclusions:** Variance in reported z-scores due to amendments do not show systematic patterns that may be attributed to modeling updates. On the other hand, they appear to have some association with COVID-19 pandemic events. Full publication of data and models should clarify the sources of non-persistence and in the published reports.

## 1. Introduction

The EuroMOMO^1^ (European Mortality Monitoring) project was funded by the European Commission (DG SANCO grant 20072001) resulting in a collaborative effort to produce real time (measured in weeks) reporting on all-cause mortality at European level. Its stated goal is “to detect and measure excess deaths related to seasonal influenza, pandemics, and other public health threats”. The lessons from 2009 pandemic Nicoll et al. [2] included the need of such a monitoring tool for the “investigation of the effectiveness of countermeasures (and the safety of pharmaceutical countermeasures)”. All-cause deaths information has been used for the indirect assessment of influenza caused mortality Schmidt et al. [4] following a line of research correlating influenza like illness reporting and all-cause mortality Nielsen et al. [3] as a surrogate to estimate the actual death toll of influenza seasons.

The Open Science philosophy and practices Bertram et al. [1] that has been embraced by the European Commission and is mandated for currently funded research projects implies that data and tools (computer code) should be open access in order to allow for science acceleration as well as independent validation of claimed results, i.e. reproducibility becoming a key concept in data science. Hence, this study aims at a retrospective systematic examination of the variability of the all-cause all-ages mortality z-scores published in the EuroMOMO site since the data download was facilitated for the first time (November 18, 2021) due to amendments that have not been explained anywhere.

## 2. Methods

Data have been extracted from stored versions of the web site found in the Internet Archive^2^. Since November 18, 2021, EuroMOMO data download was allowed. We have accessed the site versions stored in the Internet Archive the first day of each month since December 2021 until October 2023, downloading a total of 25 EuroMOMO data publication instances. Each instance consists of the the aggregated all-causes all-ages mortality z-scores for each reporting country from January 2018 until the date of the publication. The last three weeks of each instance are discarded as they are labeled as highly uncertain by EuroMOMO due to reporting delays. For each country and each week from January 2018 up to October 2023, the variance of the reported values is computed across all downloaded versions of the published data. Later weeks that are not present in earlier time series samples are treated as NaN and ignored for these computations. All the data, and python code is already published in the zenodo repository^3^.

## 3. Results

Figure 2 contains the plots of the weekly variances due to amendments of published all-causes all-ages mortality z-scores for most countries reported in the EuroMOMO. We have excluded countries with negligible variability for reasons of space. A Pooisson distribution was fitted to the pooled variances values (*λ* = 0.1437, 95%CI [0.1352, 01522]). Weeks of significant variance (*p <* 0.01) are identified for almost all countries. The list (number of weeks within brackets) is as follows : France (37), Norther Ireland (34), UK-England (31), Germay-Berlin (22), Germany-Hesse (9), Greece and UK-Scotland (6), Italy and Spain (5), Ireland and Italy (4), Germany, Hungary, Portugal, and UK-Wales (3), and Netherlans (2). Weeks of extreme variability (*p <* 0.01) happen in Germany-Berlin (14), UK-England (12), France (9), UK-Nothern Ireland (8), Germanny-Hesse (5), Hungary, Ireland, Spain, UK-Scotland, and UK-Wales (2), and Germany, Italy, and Israel (1).

**Figure 1.**
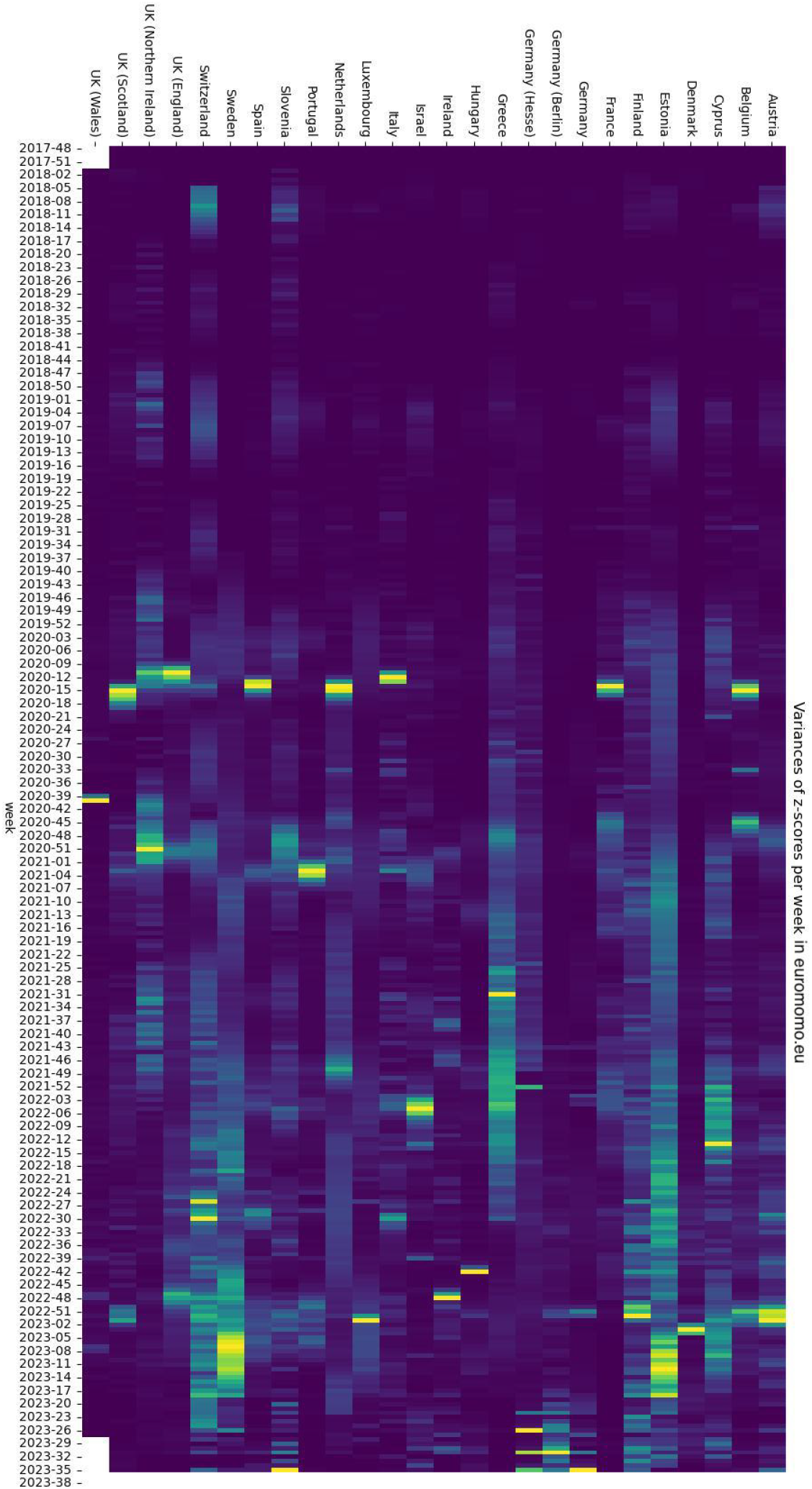
Heat map of the Variances of z-scores in Euromomo

**Figure 2.**
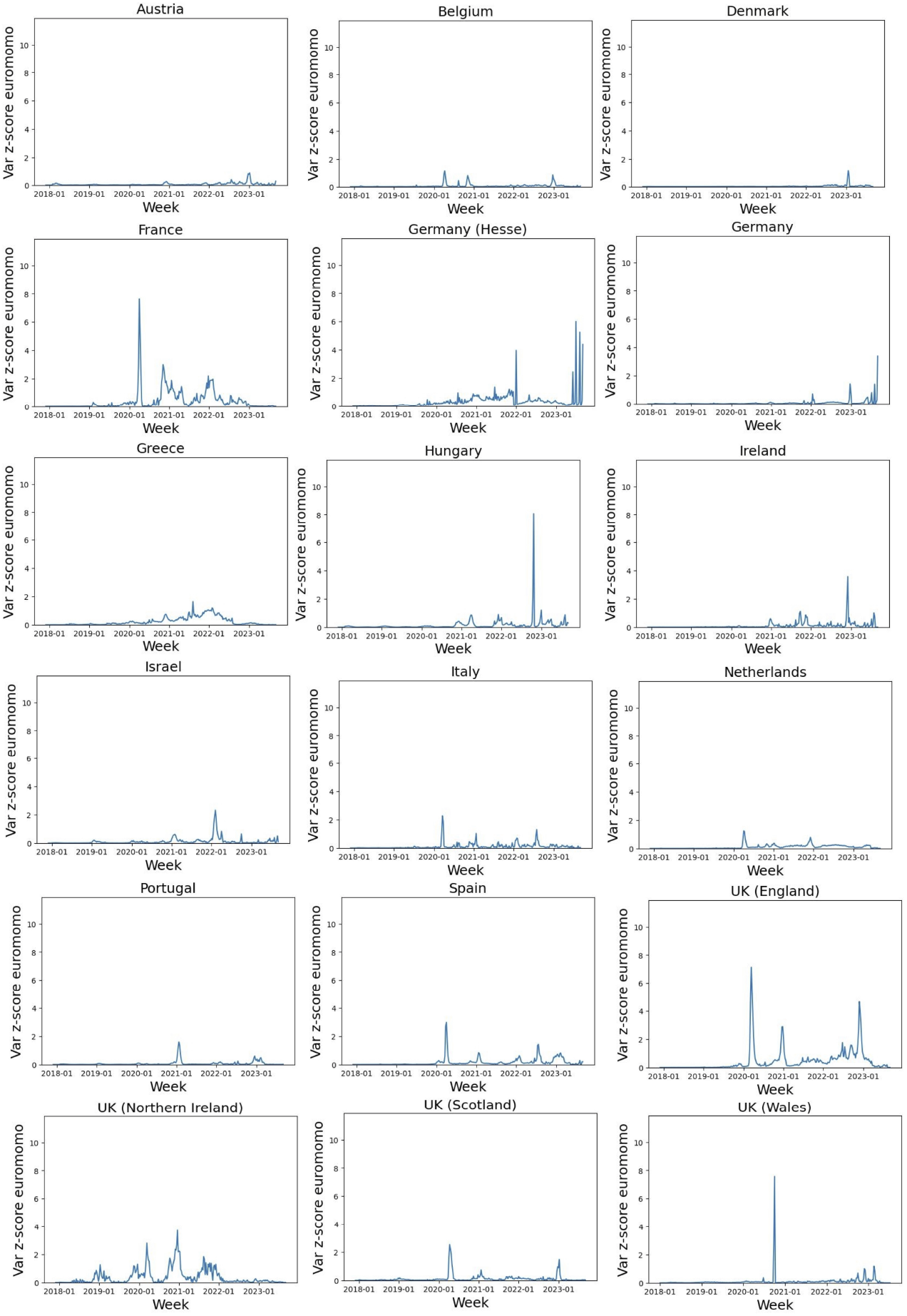
Plots of the weekly variance due to amendments of all-causes and all-ages mortality z-scores reports of countries included in EuroMOMO reporting. Variances are computed accross the monthly reports recovered from Web Archive.

## 4. Discussion

Pre-pandemic reporting significant variability was only found in UK North Ireland in the year 2019. Significant (p<0.01) variances in reported z-scores seem to be concentrated at specific periods that have some relation with events in the pandemic evolution:

- The spring of 2020 (significant variances found in weeks 9 to 16 of 2020 in France, Spain, Italy, Netherlands, UK England, Northern Ireland, and Scotland) corresponds to the deadly first wave of the COVID-19 pandemic in these countries, some of whom reported the biggest COVID-19 related mortality accross the world in this period of time. Since the first data downloads by the euromomo.eu managers were allowed at November 18, 2021, these findings mean that reported z-scores were subject to changes several years after the consolidated publication.
- The autumn and end of year 2020 (significant reporting variances found in weeks 39 to 49 of 2020 in France, Greece, Ireland, Italy, Netherlands, UK-England, UK-Wales, and UK-North Ireland) brought a second wave of COVID-19 mortality with associated increased all-cause mortality.
- At the beginning of year 2021 (significant reporting variances found in weeks 0 to 4 of 2021 in UK-England, UK-North Ireland, and UK-Scotland, Spain, Portugal, Netherlands, Italy, Israel, Hungary, Greece, Germany-Hesse, and France) most western European countries started the mass-vaccination campaign, while UK started the COVID-19 mass vaccination campaign in December 2020.
- The autumn of 2021 (significant reporting variances found in weeks 39 to 49 of 2021 in France, Germany-Hesse, Greece, Hungary, Ireland, Netherlands, UK-England and UK-North Ireland) corresponds to the vaccination of the younger populations and the booster campaigns in Europe.
- The beginning of 2022 (significant reporting variances found in weeks 0 to 5 of 2022 in France, Greece, Israel, UK-England, UK-Norther) corresponds to the explosion of COVID-19 cases in the so-called Omicron wave.
- The autumn of 2022 up to the end of the observation (significant reporting variances found in Hungary, Ireland, Spain, Germany, Germany-Berlin, Germany-Hesse, UK-England, UK-Wales) corresponds to the waning period of the pandemic, with countries stoping tests, vaccination efforts and non-pharmaceutical interventions, with reports of non-decreasing excess deaths entering the conversation.

### Limitations

The strategy of time sampling in the Internet Archive misses changes that are outlived during the month. A more comprehensive effort should revisit each and every day in order not to miss these changes, as well as the data at disaggregated age levels.

## 5. Conclusions

The results suggest that the amendments to the reported all-cause all-ages mortality z-scores do not follow a pattern consistent with changes in the regression model or the systematic upgrade of reporting systems. Furthermore, the study finds amendments on reporting of spring 2020 happening after December 2021. It is difficult to guess why after more than one year the reporting of all-cause all-age mortality can need amendments in specific countries. However, we find that significant variances in reporting due amendments are closely related to critical periods of the COIVD-19 pandemic.

In the accessed literature on EuroMOMO methods, there are little precisions about the regresion model, and there is not published code as far as the searches carried out for this study have been able to dig out. As long as the EuroMOMO reporting can be used as a tool to shape decision making in public health, it seems in the public interest that the data and the computer code are fully accessible to achieve desirable transparency for a democratic public health management.

## Data Availability

https://zenodo.org/doi/10.5281/zenodo.12716222

http://web.archive.org

## Author statements

### Ethics

the study did not required any ethical validation because it is only concerned with published data.

## Footnotes

1 https://euromomo.eu

2 http://web.archive.org

3 https://zenodo.org/doi/10.5281/zenodo.12716222

